# Use of dialysis, tracheostomy, and extracorporeal membrane oxygenation among 842,928 patients hospitalized with COVID-19 in the United States

**DOI:** 10.1101/2020.11.25.20229088

**Authors:** Edward Burn, Anthony G. Sena, Albert Prats-Uribe, Matthew Spotnitz, Scott DuVall, Kristine E. Lynch, Michael E. Matheny, Fredrik Nyberg, Waheed-Ul-Rahman Ahmed, Osaid Alser, Heba Alghoul, Thamir Alshammari, Lin Zhang, Paula Casajust, Carlos Areia, Karishma Shah, Christian Reich, Clair Blacketer, Alan Andryc, Stephen Fortin, Karthik Natarajan, Mengchun Gong, Asieh Golozar, Daniel Morales, Peter Rijnbeek, Vignesh Subbian, Elena Roel, Martina Recalde, Jennifer C.E. Lane, David Vizcaya, Jose D. Posada, Nigam H. Shah, Jitendra Jonnagaddala, Lana Yin Hui Lai, Francesc Xavier Avilés-Jurado, George Hripcsak, Marc A. Suchard, Otavio T. Ranzani, Patrick Ryan, Daniel Prieto-Alhambra, Kristin Kostka, Talita Duarte-Salles

**Author notes:** joint first authors (Edward Burn, Anthony G. Sena). joint senior authors (Kristin Kostka, Talita Duarte-Salles).

## Abstract

**Objective:** To estimate the proportion of patients hospitalized with COVID-19 who undergo dialysis, tracheostomy, and extracorporeal membrane oxygenation (ECMO).

**Design:** A network cohort study.

**Setting:** Seven databases from the United States containing routinely-collected patient data: HealthVerity, Premier, IQVIA Hospital CDM, IQVIA Open Claims, Optum EHR, Optum SES, and VA-OMOP.

**Patients:** Patients hospitalized with a clinical diagnosis or a positive test result for COVID-19.

**Interventions:** Dialysis, tracheostomy, and ECMO.

**Measurements and Main Results:** 842,928 patients hospitalized with COVID-19 were included (22,887 from HealthVerity, 77,853 from IQVIA Hospital CDM, 533,997 from IQVIA Open Claims, 36,717 from Optum EHR, 4,336 from OPTUM SES, 156,187 from Premier, and 10,951 from VA-OMOP). Across the six databases, 35,192 (4.17% [95% CI: 4.13% to 4.22%]) patients received dialysis, 6,950 (0.82% [0.81% to 0.84%]) had a tracheostomy, and 1,568 (0.19% [95% CI: 0.18% to 0.20%]) patients underwent ECMO over the 30 days following hospitalization. Use of ECMO was more common among patients who were younger, male, and with fewer comorbidities. Tracheostomy was broadly used for a similar proportion of patients regardless of age, sex, or comorbidity. While dialysis was generally used for a similar proportion among younger and older patients, it was more frequent among male patients and among those with chronic kidney disease.

**Conclusion:** Use of dialysis among those hospitalized with COVID-19 is high at around 4%. Although less than one percent of patients undergo tracheostomy and ECMO, the absolute numbers of patients who have undergone these interventions is substantial.

## Background

Treatment of patients hospitalized with coronavirus disease 2019 (COVID-19) may involve a range of medical interventions. Three distinct invasive interventions that can be readily identified in routinely collected health data and have been used in the treatment of severe COVID-19 are extracorporeal membrane oxygenation (ECMO), tracheostomy, and dialysis. For those patients with refractory hypoxemia, ECMO provides an advanced organ support alternative.^1^ Tracheostomy gives a means of facilitating long-term mechanical ventilation among critically ill patients.^2^ Additionally, the wide-ranging effects of COVID-19 are also seen with the need for dialysis to support kidney function among patients with an acute kidney injury.^3,4^ There remains uncertainty around the optimal use of each of these interventions among patients with COVID-19. For those patients who do undergo them, the use of these interventions can be taken to indicate severe disease and, for survivors, will likely be associated with long-term morbidity.^1,5–7^

Evidence on the extent of the use of invasive interventions among individuals hospitalized with COVID-19 can help improve our understanding of patient outcomes, inform healthcare resource planning, and provide an indication of some of the long-term consequences of the disease. Our objective in this study was therefore to describe the use of ECMO, tracheostomy, and dialysis among patients hospitalized with COVID-19.

## Methods

Seven large databases containing routinely-collected health care data from the United States (US) provided the basis for the analysis, with each mapped to the Observational Medical Outcomes Partnership Common Data Model (OMOP CDM). The HealthVerity database contains information on individuals with a test for COVID-19 with linkage to medical claims and pharmacy data. The Premier Healthcare Database (Premier) includes clinical coding, hospital cost, and patient billing data. The IQVIA Hospital charge data masters (CDM) includes data from resource management software within short-term, acute-care and non-federal hospitals, while IQVIA Open Claims captures open, pre-adjudicated medical claims. Optum® de-identified COVID-19 Electronic Health Record dataset (Optum EHR) Dataset represents Optum’s EHR data, while Optum® De-Identified Clinformatics® Data Mart Database – Socio-Economic Status Database (Optum SES) is an adjudicated administrative health claims database. The Department of Veterans Affairs OMOP (VA-OMOP) database reflects the national Department of Veterans Affairs health care system. This study is part of the ongoing Observational Health Data Sciences and Informatics (OHDSI) Characterizing Health Associated Risks, and Your Baseline Disease In SARS-COV-2 (CHARYBDIS) project, with the findings presented here based on data submitted as of 1^st^ October 2020.

Patients hospitalized with COVID-19 were identified in the same way as in a previous study using the OMOP CDM.^8^ COVID-19 hospitalizations ran up to March 2020 in OPTUM SES, June 2020 in HEALTHVERITY and VA-OMOP, July 2020 in IQVIA Hospital CDM, September in Premier, and October 2020 in IQVIA Open Claims and OPTUM EHR. The characteristics of study participants up to and including each individuals’ date of hospitalization (index date) were extracted, including age, sex and comorbidities (asthma, autoimmune condition, chronic kidney disease [CKD], chronic obstructive pulmonary disease [COPD], type 2 diabetes, hypertension, and obesity).

Instances of ECMO, tracheostomy, and dialysis were identified between the index date and up to 30 days following the date. Instances of dialysis were also identified in the interval from 30 days to 1 day prior to index date. The proportion of patients who underwent the interventions was calculated for each database, and stratified by age (65 or younger, and over 65), sex, and comorbidities of interest.

The entire list of definitions used to identify patients with a COVID-19 hospitalization, their comorbidities, and interventions of interest can be explored at https://github.com/ohdsi-studies/Covid19CharacterizationCharybdis/blob/master/documents/CharybdisPhenotypeLibrary.csv.

## Results

A total of 842,928 patients hospitalized with COVID-19 were included. Their baseline characteristics are described in Table 1.

**Table 1.**
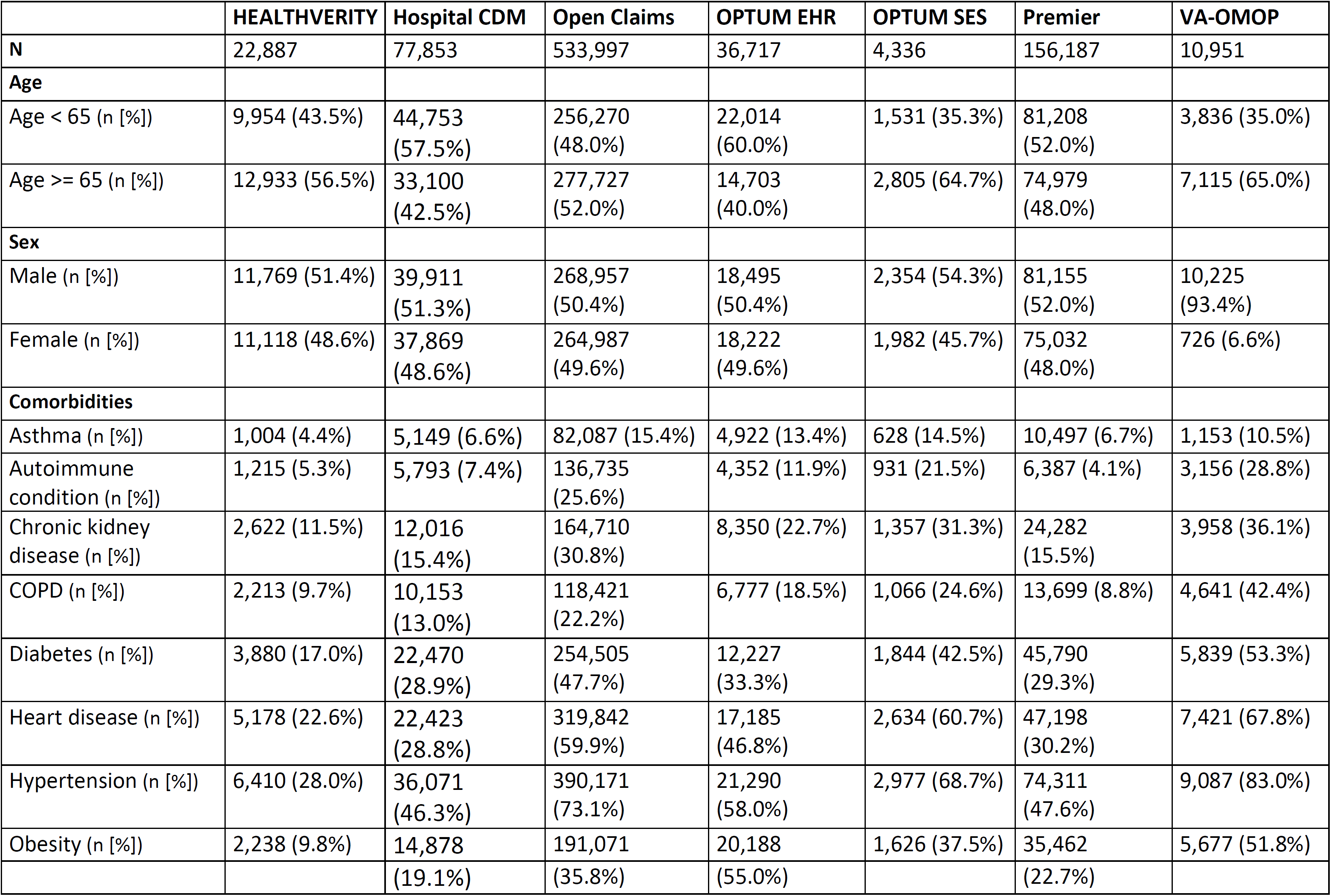
Characteristics of study cohorts in included databases.

|  | HEALTHVERITY | Hospital CDM | Open Claims | OPTUM EHR | OPTUM SES | Premier | VA-OMOP |
| --- | --- | --- | --- | --- | --- | --- | --- |
| **N** | 22,887 | 77,853 | 533,997 | 36,717 | 4,336 | 156,187 | 10,951 |
| **Age** |  |  |  |  |  |  |  |
| Age < 65 (n [%]) | 9,954 (43.5%) | 44,753 (57.5%) | 256,270 (48.0%) | 22,014 (60.0%) | 1,531 (35.3%) | 81,208 (52.0%) | 3,836 (35.0%) |
| Age >= 65 (n [%]) | 12,933 (56.5%) | 33,100 (42.5%) | 277,727 (52.0%) | 14,703 (40.0%) | 2,805 (64.7%) | 74,979 (48.0%) | 7,115 (65.0%) |
| **Sex** |  |  |  |  |  |  |  |
| Male (n [%]) | 11,769 (51.4%) | 39,911 (51.3%) | 268,957 (50.4%) | 18,495 (50.4%) | 2,354 (54.3%) | 81,155 (52.0%) | 10,225 (93.4%) |
| Female (n [%]) | 11,118 (48.6%) | 37,869 (48.6%) | 264,987 (49.6%) | 18,222 (49.6%) | 1,982 (45.7%) | 75,032 (48.0%) | 726 (6.6%) |
| **Comorbidities** |  |  |  |  |  |  |  |
| Asthma (n [%]) | 1,004 (4.4%) | 5,149 (6.6%) | 82,087 (15.4%) | 4,922 (13.4%) | 628 (14.5%) | 10,497 (6.7%) | 1,153 (10.5%) |
| Autoimmune condition (n [%]) | 1,215 (5.3%) | 5,793 (7.4%) | 136,735 (25.6%) | 4,352 (11.9%) | 931 (21.5%) | 6,387 (4.1%) | 3,156 (28.8%) |
| Chronic kidney disease (n [%]) | 2,622 (11.5%) | 12,016 (15.4%) | 164,710 (30.8%) | 8,350 (22.7%) | 1,357 (31.3%) | 24,282 (15.5%) | 3,958 (36.1%) |
| COPD (n [%]) | 2,213 (9.7%) | 10,153 (13.0%) | 118,421 (22.2%) | 6,777 (18.5%) | 1,066 (24.6%) | 13,699 (8.8%) | 4,641 (42.4%) |
| Diabetes (n [%]) | 3,880 (17.0%) | 22,470 (28.9%) | 254,505 (47.7%) | 12,227 (33.3%) | 1,844 (42.5%) | 45,790 (29.3%) | 5,839 (53.3%) |
| Heart disease (n [%]) | 5,178 (22.6%) | 22,423 (28.8%) | 319,842 (59.9%) | 17,185 (46.8%) | 2,634 (60.7%) | 47,198 (30.2%) | 7,421 (67.8%) |
| Hypertension (n [%]) | 6,410 (28.0%) | 36,071 (46.3%) | 390,171 (73.1%) | 21,290 (58.0%) | 2,977 (68.7%) | 74,311 (47.6%) | 9,087 (83.0%) |
| Obesity (n [%]) | 2,238 (9.8%) | 14,878 | 191,071 | 20,188 | 1,626 (37.5%) | 35,462 | 5,677 (51.8%) |
|  |  | (19.1%) | (35.8%) | (55.0%) |  | (22.7%) |  |

Across the seven databases, 1,568 (0.19% [95% CI: 0.18% to 0.20%)] patients underwent ECMO. The proportion of patients who underwent ECMO ranged from 0.10% (0.06% to 0.15%) in HealthVerity to as high as 0.26% (0.20% to 0.31%) in OPTUM EHR. ECMO was more often seen among patients under 65 and males, Figure 1. In IQVIA Open Claims, for example, 0.33% (0.30% to 0.35%) of those under 65 underwent ECMO while only 0.03% (0.02% to 0.03%) of those 65 or older did so, and 0.22% (0.21% to 0.24%) of men received the intervention compared to 0.12% (0.10% to 0.13%) of women. ECMO was generally used less for most of the comorbidities considered (Figure 2).

**Figure 1.**
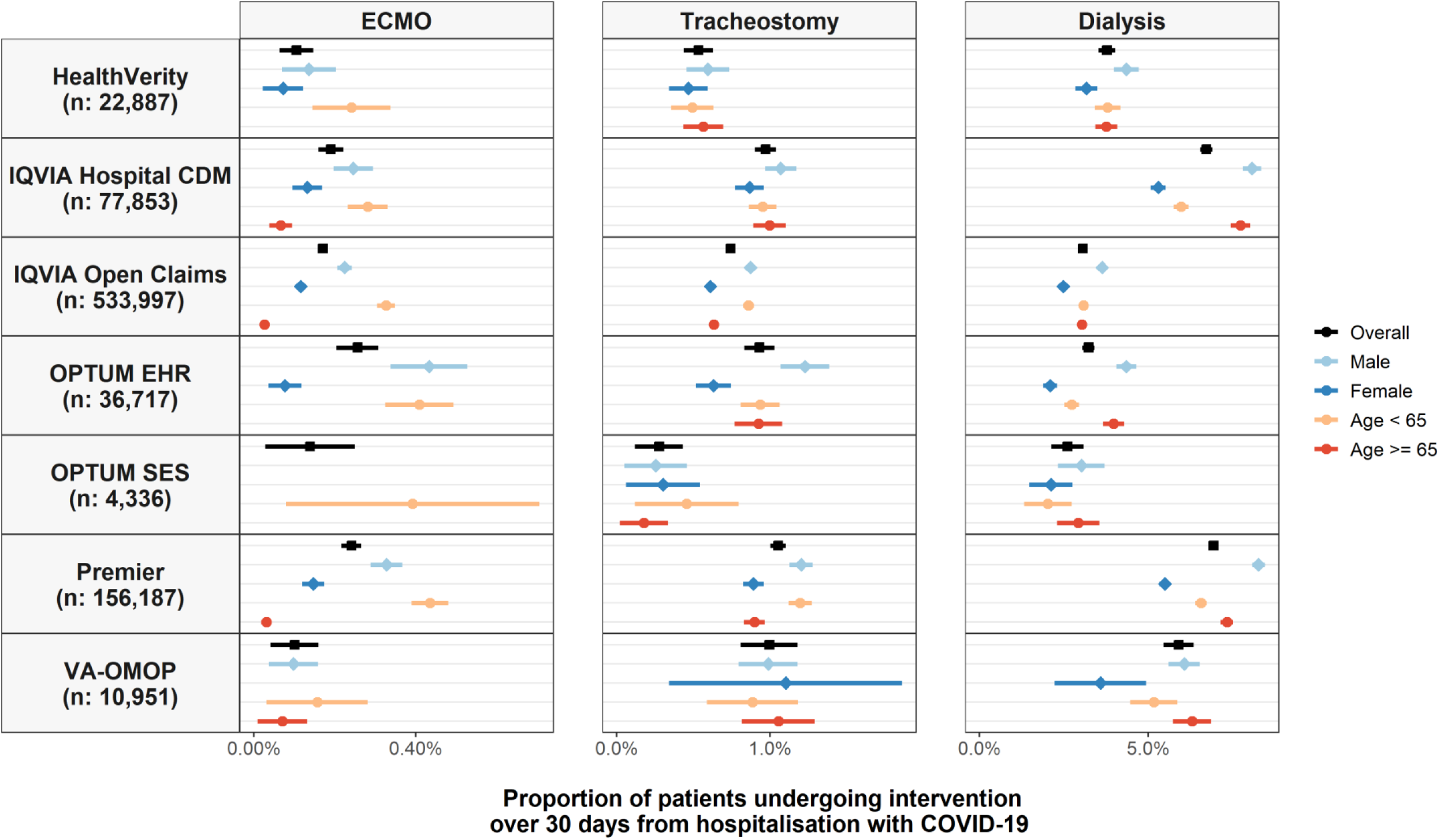
Proportion of patients hospitalized with COVID-19 who underwent ECMO, tracheostomy, or dialysis, overall and stratified by age and sex. Point estimates with 95% confidence intervals (counts of less than 10 have been omitted).

**Figure 2.**
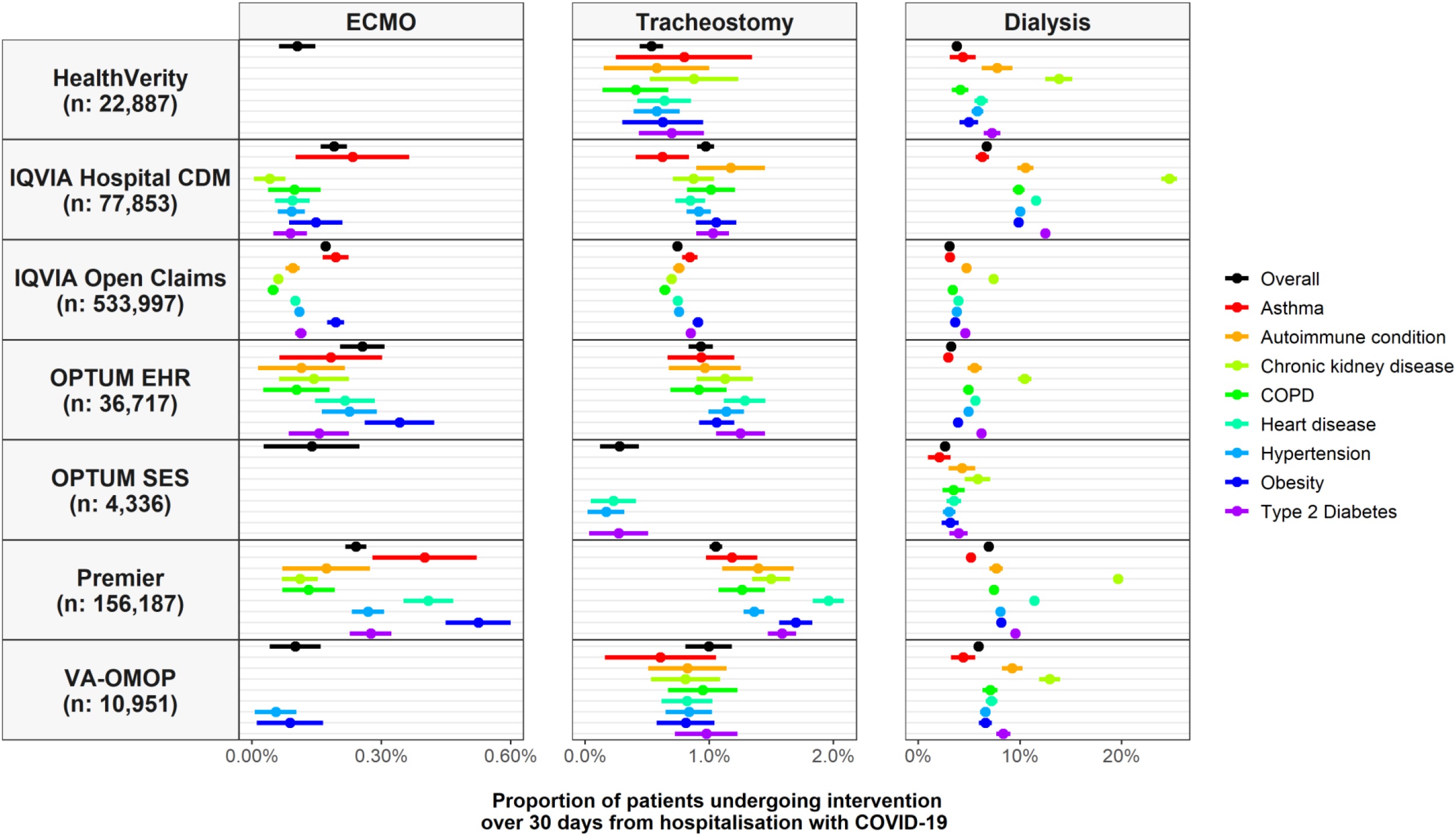
Proportion of patients hospitalized with COVID-19 who underwent ECMO, tracheostomy, or dialysis, overall and stratified by comorbidities of interest. Point estimates with 95% confidence intervals (counts of less than 10 have been omitted).

In total, 6950 (0.82% [0.81% to 0.84%]) had a tracheostomy, with this proportion ranging from 0.28% (0.12% to 0.43%) in OPTUM SES to 1.05% (1.00% to 1.10%) in Premier. Use of tracheostomy was broadly similar by age and sex, Figure 1, and by comorbidity, Figure 2, across databases.

A total of 35,192 (4.17% [4.13% to 4.22%]) patients received dialysis over the 30 days following hospitalization across the six databases, ranging from 2.61% (2.13% to 3.08%) in OPTUM SES to 6.93% (6.80% to 7.06%) in Premier. In comparison, in the 30 days prior to hospitalization, 0.6% of patients in VA-OMOP, 0.5% in OPTUM EHR, and 1.5% in IQVIA Open Claims were seen to have undergone dialysis. Use of dialysis was similar by age but was more common among men, Figure 1. In IQVIA Open Claims, for example, 3.6% (3.6% to 3.7%) of men underwent dialysis while 2.5% (2.4% to 2.5%) of women did so. Dialysis was more common among those with pre-existing CKD, Figure 2. In Premier, 19.6% (19.1% to 20.1%) of those with CKD underwent dialysis after being hospitalized compared to 4.6% (4.5% to 4.7%) for those without. Dialysis was also typically slightly more common for various other comorbidities (Figure 2).

## Discussion

Prior to COVID-19, ECMO, tracheostomy, and dialysis for acute renal failure have each been associated with poor long-term health outcomes.^1,5–7^ While continued follow-up is required to observe the long-term outcomes of the patients hospitalized with COVID-19 and underwent such interventions, it can be expected that those individuals that survived their hospitalization will face long-term morbidity and require ongoing care.

Around 4% of patients hospitalized with COVID-19 identified in this study were seen to have undergone dialysis between the day of their admission and 30 days later. This is broadly in line with the findings of a multi-center study across 12 hospitals in New York City, where around 4% of 5,700 patients hospitalized with COVID-19 received kidney replacement therapy.^9^ Higher use has though been seen for the Mount Sinai hospital system in New York City, where 9% of 3,993 hospitalized patients underwent dialysis,^10^ and among the first 1,000 patients hospitalized in the NewYork-Presbyterian/Columbia University Irving Medical Center, where 14% received dialysis.^11^ These higher rates might reflect differences in patient populations, if those admitted in these centers had particularly severe disease, or differences in treatment protocols. Given their rare occurrence, relatively few studies have reported on the use of tracheostomy and ECMO among COVID-19 patients. One of the aforementioned studies showed the use of ECMO to be 0.6%,^11^ which is slightly higher than seen in this study, which likely reflects both the setting and the severity of the patients seen at this center.

In this study we have focused on three invasive interventions which could be readily identified in each of the databases. The consistency of our results across databases is reassuring. However, it is possible that use of interventions may be underestimated if not all interventions used are unambiguously reported. This, for example, may be the concern for dialysis which when performed in intensive care may not have been recorded with a specific code, but rather billed as part of overall intensive care stay. In addition, among the databases with limited prior observation time, comorbidities can also be expected to be underreported, as can be seen with obesity in particular in HEALTHVERITY and Premier. Further interventions are of interest but were beyond the scope of this study. In particular, while we also assessed the feasibility of summarizing the use of mechanical ventilation, this was seen to have heterogeneous reporting across databases.

The findings from this study underline the severity of disease among hospitalized individuals with COVID-19. The long-term consequences for individuals who underwent the interventions and were discharged alive are likely to be substantial, both in terms of morbidity for the patients and economic burden for both patients and the health system, with individuals likely to have long-term health care needs.

## Data Availability

Open Science is a guiding principle within OHDSI. As such, we provide unfettered access to all open-source analysis tools employed in this study via https://github.com/OHDSI/, as well as all data and results artefacts that do not include patient-level health information via https://data.ohdsi.org/Covid19CharacterizationCharybdis/. Data partners contributing to this study remain custodians of their individual patient-level health information and hold either IRB exemption or approval for participation."

https://github.com/OHDSI/

## Ethical approvals

All the data partners received Institutional Review Board (IRB) approval or exemption. The use of VA data was reviewed by the Department of Veterans Affairs Central Institutional Review Board (IRB) and was determined to meet the criteria for exemption under Exemption Category 4(3) and approved the request for Waiver of HIPAA Authorization. Other databases used (HealthVerity, Premier, IQVIA Open Claims, Optum EHR, and Optum SES) are commercially available, syndicated data assets that are licensed by contributing authors for observational research. These assets are de-identified commercially available data products that could be purchased and licensed by any researcher. The collection and de-identification of these data assets is a process that is commercial intellectual property and not privileged to the data licensees and the co-authors on this study. Licensees of these data have signed Data Use Agreements with the data vendors which detail the usage protocols for running retrospective research on these databases. All analyses performed in this study were in accordance with Data Use Agreement terms as specified by the data owners. As these data are deemed commercial assets, there is no Institutional Review Board applicable to the usage and dissemination of these result sets or required registration of the protocol with additional ethics oversight. Compliance with Data Use Agreement terms, which stipulate how these data can be used and for what purpose, is sufficient for these commercial entities. Further inquiry related to the governance oversight of these assets can be made with the respective commercial entities: IQVIA (iqvia.com) and Optum (optum.com). At no point in the course of this study were the authors of this study exposed to identified patient-level data. All result sets represent aggregate, de-identified data that are represented at a minimum cell size of >5 to reduce potential for re-identification.

